# Improved Youth Wellness and Community Engagement through a built environment initiative in Appalachian Tennessee

**DOI:** 10.1101/2025.09.18.25336103

**Authors:** Phoebe Tchoua, Fenose Osedeme, Valerie Walters, Deborah Quesenberry

## Abstract

Grounded in Coordinated School Health principles, this study evaluated the impact of enhancements to an existing school-campus walking trail on student well-being and community engagement. Using surveys and validated SOPRAC observations, we assessed perceptions of wellness and trail use among students (n=651) and staff (n=38). Most students (63%) reported that the trail positively affected their well-being, describing feeling “calm”, “energic”, “happy”, and “peaceful”. School staff (teacher n=32, staff and other, n=6) echoed these perceptions, with 88% noting moderate to strong positive impacts on student well-being and 76.5% observing improved student mindset post-trail use. Teachers described students as “refreshed,” “calm,” and “enthusiastic.” Observational data from 34 weekday and weekend trail visits showed similar usage rates between male and female students (45% vs. 43%), with adults comprising the majority of users (62%), followed by teens (30%) and children (8%). Walking was the most common activity across all groups. These findings suggest that enhancing built environments through school–community partnerships can promote physical activity and support student well-being and improved mood.

## INTRODUCTION

Low levels of physical activity (PA) among youths in the United States (U.S.) continues to be a public health concern. PA is crucial to the well-being (i.e., mood, physical activity levels) of youths as it promotes positive health outcomes, improved mental health, enhanced cognition, and weight control amongst other benefits.^1^ Studies indicate that the moods of adolescents who participate in PA significantly improved compared with the moods of those who do not participate in PA.^2, 3, 4^ Apart from helping with positive mood, PA can help adolescents to establish self-esteem.^5^ Despite the health benefits of PA, only about 1 in 4 high school students get the recommended one hour a day of PA in the U.S. placing them at an increased risk for morbidities and mortality.^6^

Appalachia is a region that spans 13 states from Northern Mississippi to Southern New York. It is characterized by increased rurality, limitations to built environment, and inconsistent PA engagement.^7,8^ Both youth and adults living in Appalachia are less physically active compared to other regions in the U.S.^9^ In rural Appalachia Kentucky, 92.38% of young adults and 97.62% of older adults do not meet the physical activity recommendation.^10^ According to self-reported data from a study in Appalachian Ohio, only 37% of high school students engaged in moderate physical activity for at least five days per week, and only 14% met the national physical activity guidelines.^11^ In Appalachian Tennessee, the rate of physical inactivity is significantly higher than national level (34.2 vs 25%).^9,12,13^

Within the State of Tennessee, there is a rural-urban divide in PA as the rate of physical inactivity within the Appalachian Tennessee (rural) region is higher than that of the non-Appalachian Tennessee (urban) regions (34.2 vs 30.6%).^9^ Additionally, research has indicated significantly lower rates of PA among Appalachian adolescents compared to national averages with the lowest rates found in rural areas as opposed to larger metro areas of the Appalachian region.^1^ Barriers to PA are discomfort, time (e.g., travel to PA facility, work, family commitments), lack of opportunity for PA, program closure due to low attendance, transportation, built environment.^14^ These conditions in Appalachian Tennessee provide an impetus for advancing evidence-based interventions promoting PA among youths in the region.

PA is a health-related behavior that has antecedents. Some of these are environmental in nature, hence the need for interventions that impact environmental and social structures.^15^ Effective strategies for promoting active lifestyles and overcoming related barriers can help increase PA levels among youths. Population-based approaches are a promising approach because they offer several benefits compared with approaches focused on individual behavior change.^16^ They are more likely to impact a greater number of people and lead to longer-lasting and favorable lifestyle changes.^17^ Previous studies show that these approaches have been recommended as an effective approach to behavior change such as PA.^16,18^

One of these population-based approaches includes built environment interventions. Built environment interventions as defined by the Community Preventive Services Task Force (CPSTF) are activities that include creating or modifying environmental characteristics in a community to make PA easier or more accessible.^19^ Evidence shows that built environment strategies may incorporate one or both of the following components: a) Pedestrian and Bicycle Transportation System Intervention, b) Land Use and Environment Design Intervention.

Findings from a systematic review by the CPSTF recommends combining more than one built environment strategy or intervention component (e.g., pedestrian and bicycle transportation, land use and environment design) to increase PA.^18^

Trails are one of the fastest growing attributes of a built environment intervention that support PA.^20^ A trail is a public path that creates an attractive transportation and leisure activity corridor for walking, running, and cycling;^21^ it supports leisure-time PA and/or active transportation. Study findings show that building trails within school campuses have multiple benefits to teachers, youths, and their families. These benefits include enabling outdoor PA by providing the environment for physical exercise.^22^

Evidence on the impact of built environment approaches in increasing PA among youth in rural communities such as Appalachian Tennessee is scarce. Our objective is to present findings from our study evaluating how grant-funded built environment improvements in a rural Northeastern Tennessee county are affecting physical and mood outcomes among students, staff, and community members. In response to concerns over the health of the youth in the state, Tennessee is unique in its integration of Coordinated School Health (CSH) programming through legislative support for its inclusion in all school districts in the state.^23^ Central to this program is the oversight provided by CSH Coordinators in each district. The enhancements to the greenway that is the subject of this investigation came about through community-school collaborations spearheaded by the CSH Coordinator (3^rd^ author) in partnership with the representative from the funding agency.

Using Coordinated School Health concepts as the foundation of the work, we sought to assess the impact of enhancement to an existing walking trail on a school campus in order to enhance student well-being and community engagement with the school environment.

## METHODS

The study used a quantitative design for the evaluation of a walking trail located between a middle and high school campus in a rural area, consisting of direct observations of trail use and self-administered electronic surveys between March 2022 until April 2023. This manuscript was written in alignment with the STROBE checklist.^24^

### Participants

Participants were community members who used the trail as well as students and staff (teacher, counselors from a regional middle and high school who completed the survey (students n=651, teachers and staff, n=38). All middle and high school students and staff (1950 high school students, 1073 middle school students, and 94 teachers and staff) were eligible to participate in the survey. This project evaluated the use of a grant-funded walking trail by students and community members. The [Blinded for review] Institutional Review Board (IRB) reviewed this project and determined it did not fall under the purview of the IRB as it was not deemed human subject research. The study collected anonymous data and obtained electronic consent from both students and staff. Participation in the survey was entirely voluntary, and individuals could opt out at any time without penalty.

### Instrumentation: Observation and Surveys

For trail observation, the team used the System for Observing Play and Recreation in Communities (SOPARC), and followed the process outlined in the SOPARC Description and Procedure Manual.^25^ SOPRAC is an observation tool designed to collect information in real-time on the use of parks by community members. During each site visit, observers completed the SOPARC Path Coding Form which asks for date, target area, park id, observer id, start and end time of each area scanned, gender (male or female), age group (child, teen, adult, senior), ethnicity (Latino, Black, White, Other), and activity level (sedentary, walking, vigorous) of each observed participant. The SOPARC protocol defines activity level as “Sedentary (S) = Individuals are lying down, sitting, or standing in place; Walking (W) = Individuals are walking at a casual pace; Vigorous (V) = Individuals are currently engaged in an activity more vigorous than an ordinary walk (e.g., increasing heart rate causing them to sweat, such as jogging, swinging, doing cart wheels)”.^26^ The data was tallied, aggregated, and transferred to the corresponding SOPARC Path Observation Form which added more details on the conditions of the target area and grouped participants by gender and activity.

The student and staff surveys consisted of 24 and 26 questions, respectively, organized into 5 main categories: demographics, trail usage, PA of students on the trail, well-being, and perceived value and satisfaction with the trail. The surveys included both open- and close-ended questions, utilizing multiple choice format, Likert-scales, and free-response items to capture a range of quantitative and qualitative data.

Students were asked about their frequency of trail use (response options: daily or most days, a few times a week, a few times a month, occasionally, or rarely), change in use following the trail redesign, and primary reason for trail use (response options: walking/running, commute to school, enjoy nature, to get away from school/life stress). PA questions focused on whether the trail contributed to increased PA, and which type of activities they could do on the trail (response options: running/jogging, walking/dog walking, biking, other activity).

Well-being items assessed the trail’s impact on their overall mood, mindset, focus in class, and emotional state while using the trail using a 5-point Likert scale from “do not feel” to “feel very strongly” for description such as refreshed, calm, fatigued, happy, etc. Finally, students shared what they enjoyed on the trail (response options: scenery, fresh air, exercise, animals, plants, insects, etc.), activities they could do now, and provided suggestions for improvements.

Staff completed a parallel version of the survey, responding from their perspective on how the trail impacted their students. Two additional items were included for the staff: their role at the school and their personal trail use.

### Observation and Survey Procedures

Data collection lasted 14 months, from March 2022 until April 2023. In all, team members collectively conducted 34 observations: one observation per trail site visit. A trail site observation visit included collecting data from a target area, usually the trailhead, where the entire trail visibility was mostly unobstructed. In order to assess trail use during various days and times, each observation round on average lasted one to one and a half hours, and took place on every day of the week (Monday to Sunday) to assess trail usage. Trail observers recorded each observation on the SOPARC Path Coding Form and the SOPARC Path Observation Form.

During the 2023 Spring semester (January - May), the CSH Coordinator contacted staff of a local Middle and High School via email and asked them to participate in the project evaluation electronic survey. In addition, physical education teachers were recruited to distribute the survey link and QR (quick response) code to their students to assess trail usage.

### Data Analysis

Descriptive statistical analyses were conducted using Microsoft Excel Version 16.72 on all data, including student and staff surveys, and observations. Descriptive analyses included frequencies and percentage of the data.

## RESULTS

### Observations

From March 2022 to April 2023, three observers conducted 34 observations encompassing each day of the week, where the majority of observations lasted one to one and a half hours. 43% of trail users were female and 45% male, and most people were adults (62%), followed by teens (30%), and lastly children (8%) (see Table 1). Across all genders, walking was the most common activity, 76% of female and 91% of male. All activities by gender are presented in Table 1.

**Table 1.** Observation – Activities on the Trail by Gender.

| □ | Female |  | Male |  | Unknown |  | Total □ |  |
| --- | --- | --- | --- | --- | --- | --- | --- | --- |
| □Activity | N | % | N | % | N | % | N | % |
| Walking | 25 | 76% | 31 | 91% | 0 | 0% | 56 | 74% |
| Sitting/Standing | 5 | 15% | 2 | 6% | 9 | 100% | 16 | 21% |
| Running | 3 | 9% | 1 | 3% | 0 | 0% | 4 | 5% |
| Total | 33 | 100% | 34 | 100% | 9 | 100% | 76 | 100% |

### Surveys

Quantitative data was collected from 651 students, where 47% were female and 45% male, and from 38 teachers and staff with 66% female and 32% male. Slightly over 1 in 4 Students reported increased trail use following trail enhancements (regraded and paved path to increase accessibility, and added lighting, disc golf course, and signage).

Thirty-five percent of teachers and staff brought their students to the trail either daily, a few times weekly or monthly. In response to the question, “In an average week, how often do you get more physical activity because of the trail?”, most students attributed some level of physical activity to the trail, and teachers and staff also reported that the trail positively impacted the students’ physical activity (see Table 2).

**Table 2.**
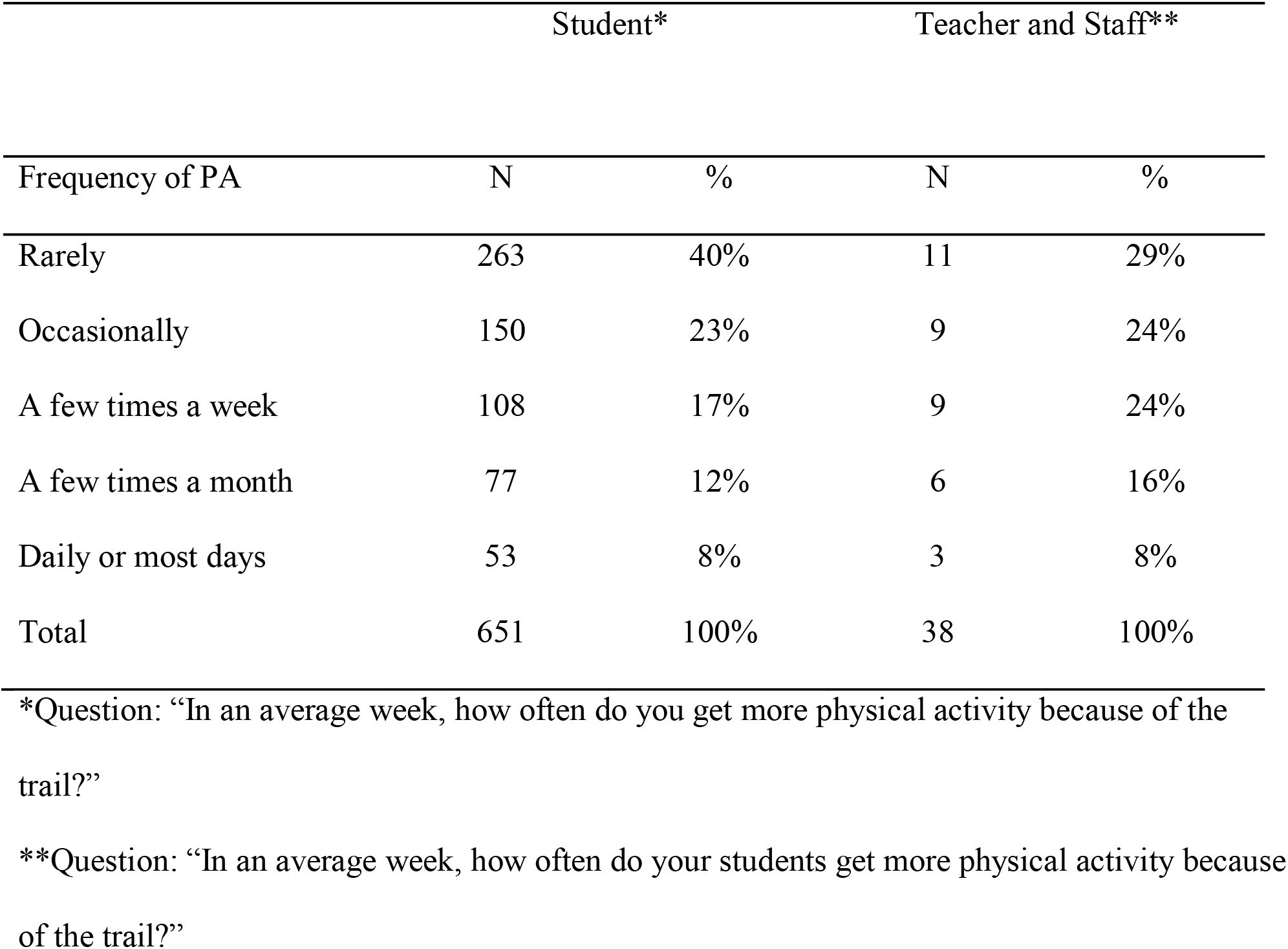
Survey – Physical Activity Because of the Trail.

Survey respondents were asked about the student activities on the trail, 71% of students and 79% of teachers and staff said, students used the trail for walking, running, exercise, or fitness. Students reported that they could run/jog (59%), walk (27%), play sports (5%), or bike (1%) as activities they are able to do on the trail.

When asked about the trail’s impact on the students’ overall well-being, 43% percent of students and 34% of teachers and staff said “moderately positive”, and 20% of students and 53% of teachers and staff said “very positive”. With regards to trail use and positive mindset, 23% and 13% of students “very often” and “always” respectively, had a positive mindset after using the trail. On the other hand, teacher perceived more students had a positive mindset.

Students were asked to indicate the extent to which 12 words described how they felt when they used the trail. Many students reported strongly feeling, happy, calm, peaceful, and refreshed. The other eight words are presented in Figure 1. When teachers were asked a similar question about how they perceived the trail’s effect on their students, many felt strongly that their students were energetic, revived, and peaceful. In response to an open-ended survey item, participants reported a wide range of activities made possible by the redesigned trail, including walking, jogging, running without falling, running on an even path, running better and longer, cross-country practice, and completing the physical education class mile run.

**Fig. 1.**
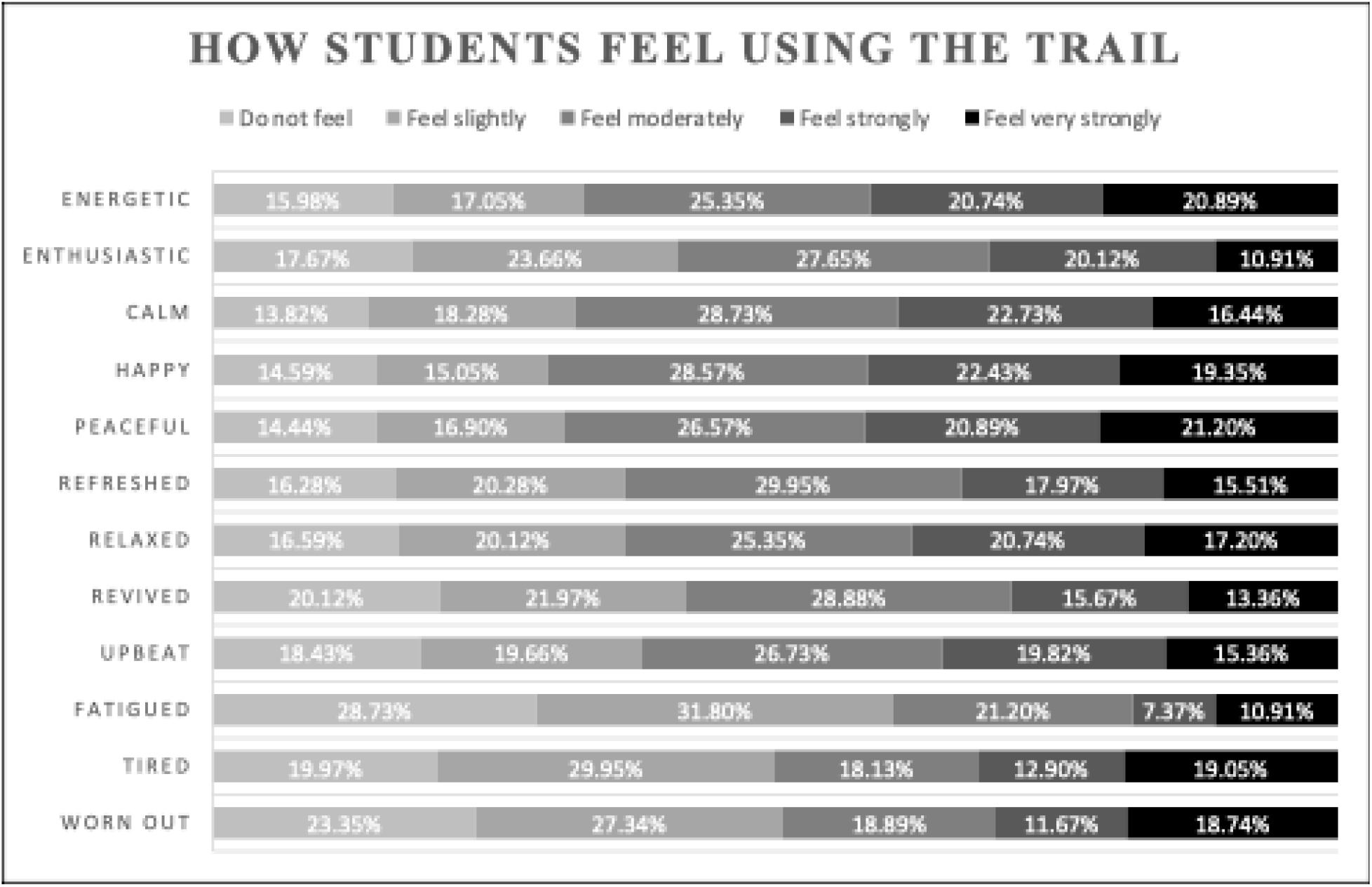
How Students Feel While Using the Trail.

## DISCUSSION

The current study presents the findings from the evaluation of the Greene County built environment initiative among youths in Appalachian Tennessee. Underserved populations such as this are of special interest due to high rates of health disparities including chronic illness and disease, poor health behaviors, and lack of health-care resources. Research on the impact of built environment in this population revealed that very little has been documented in literature related to this important topic among this population. To the best of our knowledge, we are aware of no studies that have documented the implementation of a built environment initiative on the wellness of youth in Appalachian Tennessee.

Similar to related studies, our study shows that the trail had an impact on the students’ overall well-being.^17^ In describing how youths felt after using the trail. Words such as “happy”, “calm”, “peaceful”, and “refreshed” were used. These findings are also supported by studies with evidence that built environment has positive direct and indirect effects on mood.^21,27,28^ Other studies also find that physical activity among youth also impacts moods positively. Storlaska et al^4^ found in their study of females that physical activity increased the mood of the students with the most effect on the energetic arousal of the students.

Our study used an observational design to evaluate trail usage and activities performed at the trail.^29^ This design has been used by other studies examining the impact of built environment features among young people.^30^ According to Song and Chung ^31^, study design can play an important role in deriving evidence. For our study, we had multiple researchers observe the trail at different times of the day to generate stronger evidence for the study.

Findings from this study show that implementing a neighborhood-built environment is crucial to facilitating PA among youth.

### Implications for Policy and Practice

Within schools, incorporating trails as a built environment strategy is crucial to facilitating PA, therefore increasing wellness of youths. The findings of this study have important implications for current school health practices. Given the favorable impact of the trail improvement on the overall well-being of middle and high school students in this study, policies should be implemented at the local and state level that support improvement to the built environment on school campus and encourage more physical activity outdoors for middle and high school students during the school day. School policies that encourage and support outdoor time for students beyond physical education classes might enhance positive health development of adolescents.

### Limitations

The cross-sectional nature of the study means that claims about causality are not possible. Longitudinal studies or natural experiments would add strength to the study findings. Additionally, the observational data reports observations outside school hours. The researchers were unable to gather observational data during school hours. Notwithstanding these limitations, this study addressed an existing gap in the literature and provides evidence on the impact of built environment approaches in increasing PA among youth in rural communities.

## CONCLUSIONS

The findings shared in this report support the initial goal of this grant evaluation on the built environment’s effect on the mood and well-being of community members in rural Appalachia Tennessee. The trail is clearly making a positive impact in the life of the students and employees of the rural middle and high School. The survey results show the short-term benefits of the redesigned trail, and continued survey data collection will provide the long-term benefits of this project on the community’s mood and well-being. Over time, it is anticipated that students, school staff, and community members will increase their use of the trail as they become more aware of its existence. Additionally, the partnership between a regional state university, CSH, and a Northeast Tennessee school district proved successful, and provides a framework for other schools who desire to partner with a university to improve the well-being of their students, employees, and community members. For future research, a longitudinal cohort design would be helpful to effectively understand the correlations between the trail and wellness of youths over an extended period of time.

## Human Subjects Approval Statement

The [Blinded for Review] Institutional Review Board (IRB) reviewed this evaluation project and determined it did not fall under the purview of the IRB.

## Data Availability

All data produced in the present study are available upon reasonable request to the authors

## Acknowledgments

This work was supported by the Tennessee Department of Health Project Diabetes Program. We are grateful to Ada Sloop, MPH, Community Health Program Manager, Northeast Regional Health Department of Tennessee, for her invaluable guidance throughout this study. Her insightful feedback and expertise were instrumental in shaping this research.

